# COVID-19 Vaccine Acceptance in Nigeria: A Rapid Systematic Review and Meta-Analysis

**DOI:** 10.1101/2023.02.16.23286008

**Authors:** Victory Chizaram Nnaemeka, Nnenna Audrey Okafor, Oluwatosin Qawiyy Orababar, Ruth Anikwe, Reuben Ogba Onwe, Nneka Patricia Uzochukwu, Thomas Sambo Tsiterimam, Nkiru Nenye Nwokoye, Anthony Chibuogwu Ike

## Abstract

Widespread COVID-19 vaccination is essential to maintaining pandemic control. However, low- and lower-middle-income countries (LMICs) continue to face challenges to care due to unequal access and vaccine fear despite the introduction of safe and effective immunizations. This study aimed to collect information on Nigeria’s COVID-19 vaccine uptake rates and determinants. Science Direct, PubMed, Google Scholar, African Journal Online, Springer, and Hinari were all systematically searched through and completed in May 2022. Quality assessments of the listed studies were performed using the eight-item Joanna Briggs Institute Critical Appraisal tools for cross-sectional studies. In addition, we undertook a meta-analysis to calculate pooled acceptance rates with 95% confidence intervals (CI). Forty-two studies in total satisfied the inclusion criteria and were reviewed. A total of 24,533 respondents were studied. The total sample size of states in the Northern, Western and Southern parts of Nigeria are 3,206, 4,527 and 5,059, respectively, while 11,741 is the cumulative sample size of all the Nigeria-wide studies. The total COVID-19 vaccination acceptance rate among all the study groups was 52.4% (95% CI: 46.9-57.9%, *I*^2^ = 100%), while the total estimated COVID-19 vaccination hesitancy rates was 47.81% (95% CI: 42.2 – 53.4% *I*^*2*^ = 100%). In Nigeria-regions sub-group analyses, the Western region (58.90%, 95% CI: 47.12–70.27%) and Northern region (54.9%, 95% CI: 40.11%–69.4%) showed the highest rates of vaccine acceptance and vaccine hesitancy respectively. The COVID-19 vaccine acceptance rate was highest in 2020, with a pooled rate of 59.56% (46.34, 57.32%, *I*^*2*^ = 98.7%). The acceptance rate in 2021 was only 48.48 (40.78%, 56.22%), while for the studies in 2022, it increased to 52.04% (95% CI: 35.7%, 68.15 %). The sensitization of local authorities and the dissemination of more detailed information about the COVID-19 vaccine and its safety, could significantly increase the country’s vaccination rate.

## 1. Introduction

The factors causing vaccine hesitation or acceptance are widely recognized as a complex phenomenon with various predictors beyond safety concerns (Patwary *et al*., 2022). Vaccine-hesitant individuals have been described as a heterogeneous population amid a spectrum that runs from complete acceptors to complete refusers (Dubé *et al*., 2016). These “hesitant” individuals may refuse some vaccines but agree to others, reject vaccines, or are unsure of accepting vaccines (Larson *et al*., 2014). The Strategic Advisory Group of Experts on Immunization (SAGE) identified three major factors that affect the attitude towards vaccination acceptance. This includes complacency, convenience and confidence (MacDonald & SAGE Working Group on Vaccine Hesitancy, 2015). Complacency shows a low impression of the disease risk, leading to the perception that vaccination is unnecessary (Larson *et al*., 2011). Confidence refers to the trust in vaccination safety, effectiveness, and the healthcare systems’ competence. Convenience entails the availability, affordability and delivery of vaccines in a comfortable context (Durbach, 2000). Moreover, the complex motives behind vaccine hesitancy can be analyzed using the epidemiologic triad of environment, agent and host factors (Larson *et al*., 2011; Porter & Porter, 1988). Environmental factors include public health policies, social factors and the messages spread by media (Dempsey *et al*., 2011; Gust *et al*., 2008; Robison *et al*., 2012). The agent (vaccine and disease) factors involve the perception of vaccine safety and effectiveness besides the perceived susceptibility to the disease (Gust *et al*., 2008; Luthy *et al*., 2009; Opel *et al*., 2012). Host factors depend on knowledge, previous experience, education, and income levels (Opel *et al*., 2012).

Whereas much information is known regarding vaccine acceptance in developed countries, little is known about COVID-19 vaccine acceptance in Sub-Saharan Africa (Ditekemena *et al*., 2021). Disturbingly, Nigeria once experienced a striking example of widespread vaccine refusal. This incident occurred in 2003–2004 when northern Nigeria boycotted the polio immunization program, which caused the disease to reemerge in the nation and beyond (Adebisi *et al*., 2021; Ike *et al*., 2018; Kabamba Nzaji *et al*., 2020; Tobin *et al*., 2021). LMIC residents may not be aware of the risks associated with the disease; hence, they may be less eager to undergo vaccinations as a result, given that COVID-19 mortality rates in LMICs have consistently been lower than those in higher-income nations (Brown *et al*., 2011).

A study in Nigeria found that partial immunization may be influenced by factors such as parental disapproval, maternal availability, poor knowledge, inadequate allocation of efficient vaccines, negative historical experiences involving foreign factors, cultural and religious beliefs, and mistrust of government (Danis *et al*., 2010). We do not, however, know the exhaustive classifications and confirmations of these parameters and their consequences. Earlier surveys in Nigeria have estimated vaccine acceptance and hesitancy among subgroups and the general population in different states and geopolitical zones in Nigeria. For example, studies from (Adejumo *et al*., 2021; Allagoa *et al*., 2021) estimated a vaccine acceptance of 24.60% and 53.50% among 1000 hospital patients and 1767 HCWs, respectively, in southern Nigeria states. Other studies have focused on qualitatively summarizing Nigeria’s vaccine hesitancy and acceptance rate and scoping vaccine acceptance rates in higher or lower-income countries (Aw *et al*., 2021). It has not yet been investigated whether vaccine acceptance and hesitation rates and their associated determinants exist in Nigeria using a systematic review and meta-analysis.

To determine the acceptance and reluctance rates of the COVID-19 vaccine among Nigerians, we conducted a rapid systematic review and meta-analysis. Additionally, we aimed to identify potential factors associated with vaccine acceptability in Nigeria. This study could provide the first steps for facilitating the planning of ongoing vaccination programs and enhancing vaccine uptake in developing countries as global vaccination efforts continue.

## 2. Materials and Methods

We used an expedited procedure to summarize the evidence using a rapid systematic review approach (Haby *et al*., 2016). Accordingly, the Preferred Reporting Items for Systematic Reviews and Meta-Analyses Statement (PRISMA) guidelines were followed when conducting the methodology (Page *et al*., 2021).

### 2.1. Search Strategy

A systematic search was conducted in Science Direct, PubMed, Google Scholar, African Journal Online, Springer and Hinari. Keywords in various parenthesis permutations such as “COVID-19″, “VACCINE”, “ACCEPTANCE”, and “NIGERIA”, “COVID-19 VACCINE”, “ACCEPTANCE” “NIGERIA″ “COVID-19″ “VACCINE ACCEPTANCE”, “NIGERIA″ “COVID-19 VACCINE ACCEPTANCE” “NIGERIA” “VACCINE HESITANCY” “NIGERIA WEST AFRICA” were used. Boolean operators such as (AND, NOT, OR) were also used. Finally, all the identified articles were downloaded into Zotero bibliographic management software for further processing. The last search for the studies was conducted on May 31, 2022. Further details on the search methods are described in the supplementary material.

### 2.2 Eligibility criteria for the studies

The following criteria were used to determine which studies would be included in the meta-analysis: studies containing at least one vaccine acceptance or hesitancy-related question; restricted to states and regions in Nigeria; articles published in English between January 2020 to May 2022; observational and descriptive studies with a cross-sectional, experimental, and longitudinal design. Excluded articles included those that did not attempt to assess the acceptance or hesitation of the COVID-19 vaccine, unpublished data, books, conference papers, systematic reviews, literature reviews, commentaries, editor letters, and case reports. In addition, publications without access to the full text were also excluded.

### 2.3 Study selection

After the importation of studies into the Zotero Bibliographic Software, duplications were sifted out; after that, the studies were further selected based on their titles and abstract suitability. Finally, the full text of the studies was perused to check for those that met the inclusion criteria. Publications were assessed based on the Joanna Briggs Institute (JBI) critical appraisal tool for cross-sectional studies. Each condition was awarded: 1 for yes and 0 for No/unclear/Not Available. Two independent reviewers (A.I and O.O) also reviewed the studies based on the criteria, and discrepancies in selection were resolved by discussion.

### 2.4 Data Extraction

Two reviewers extracted data. Author name, publication year, study nation, study design, survey period, target population, sampling technique, sample size, vaccine acceptance measuring scale, and factors linked to vaccine acceptance, reluctance, or refusal were all retrieved. Table 1 contains all the extracted data. Disparities were then resolved by consensus following independent data extraction.

**Table 1.**
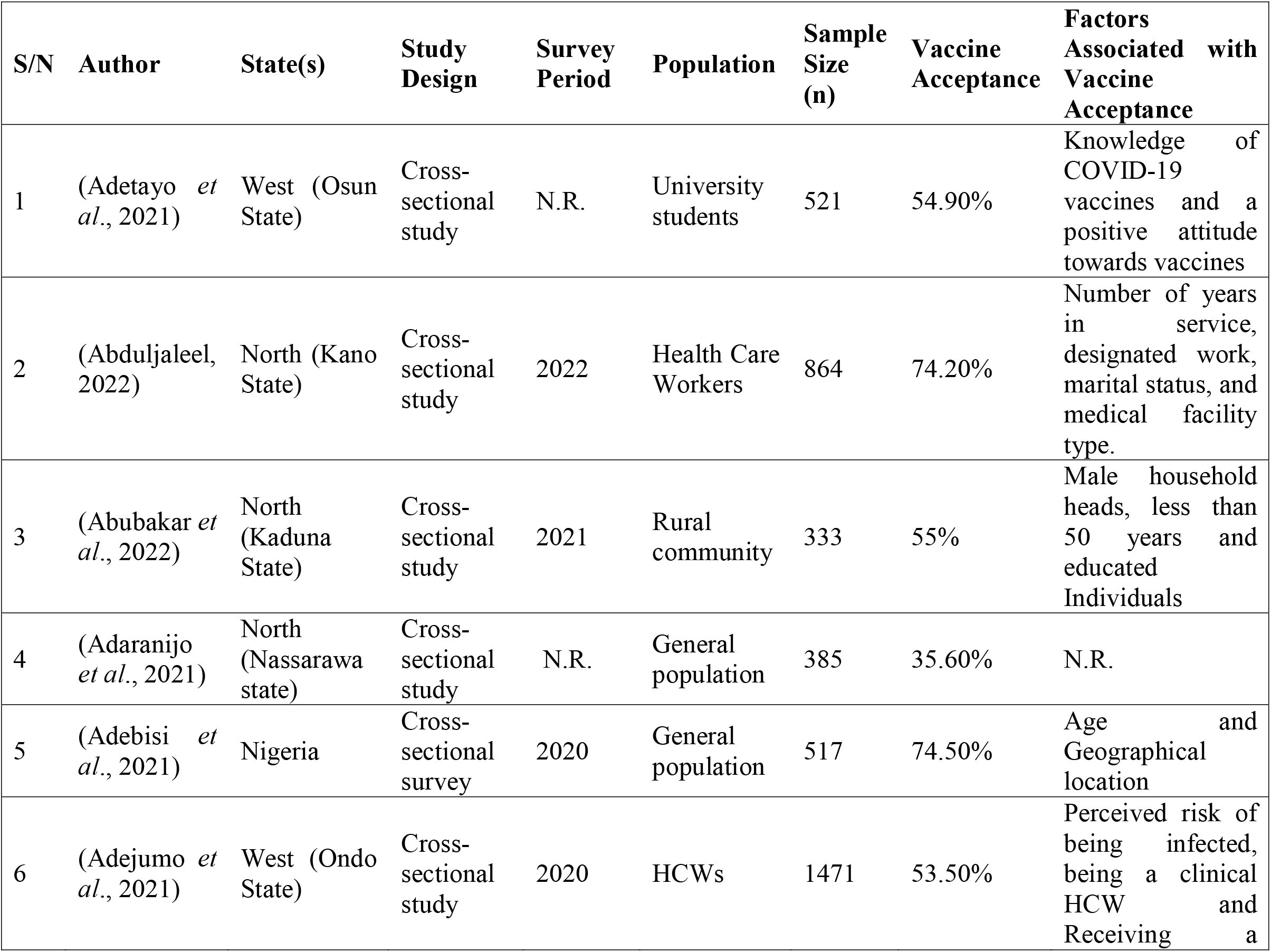
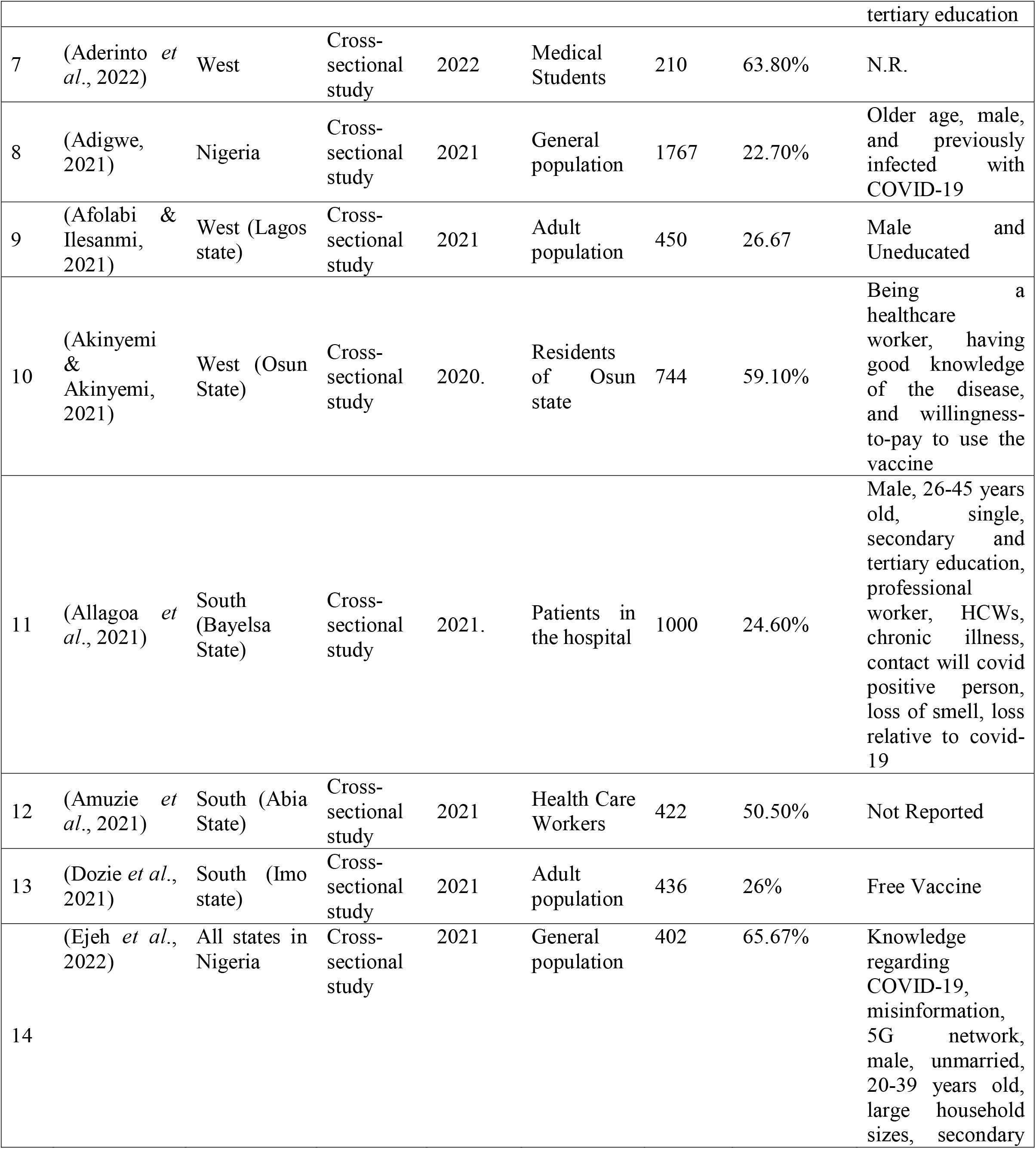
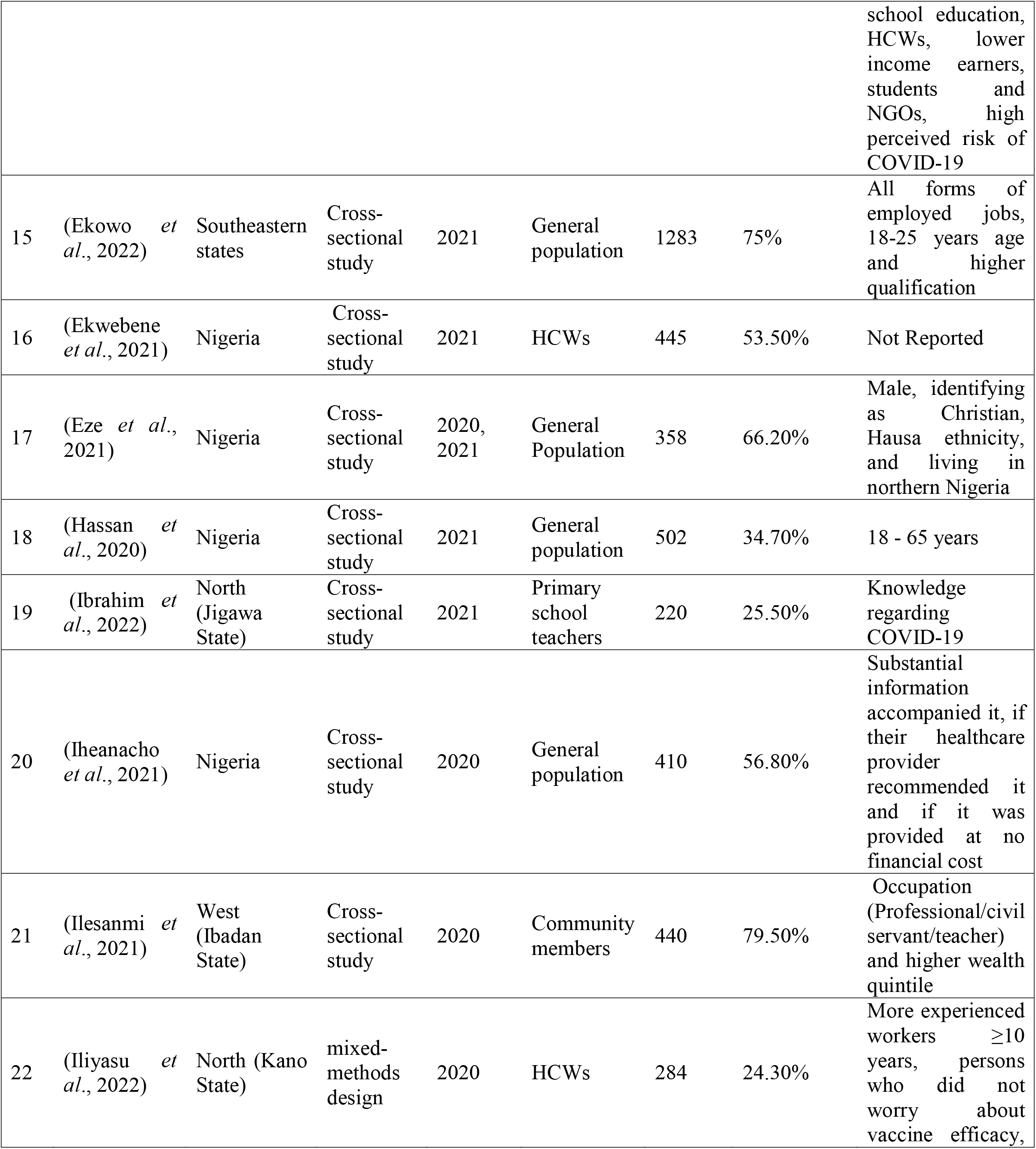
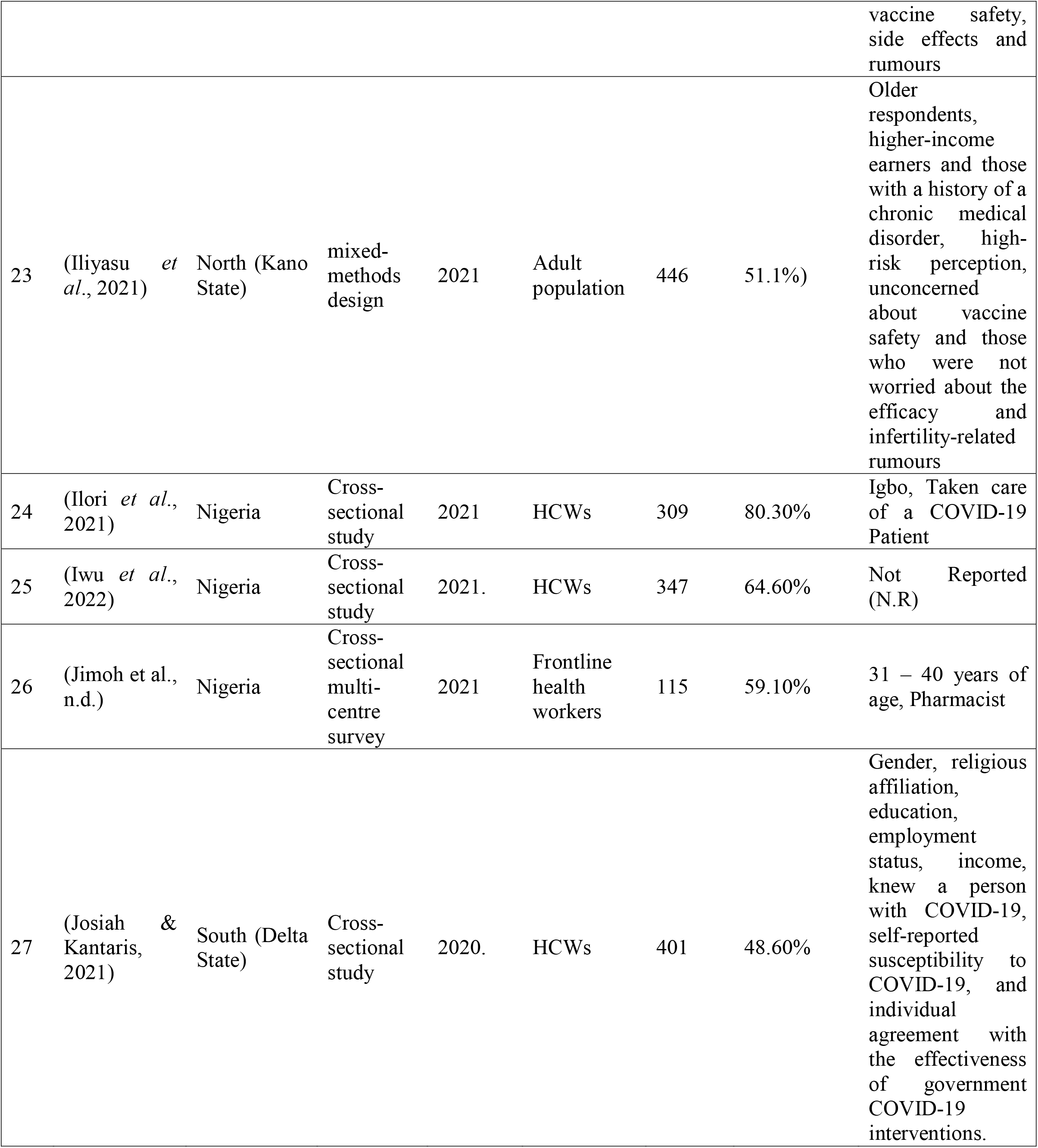
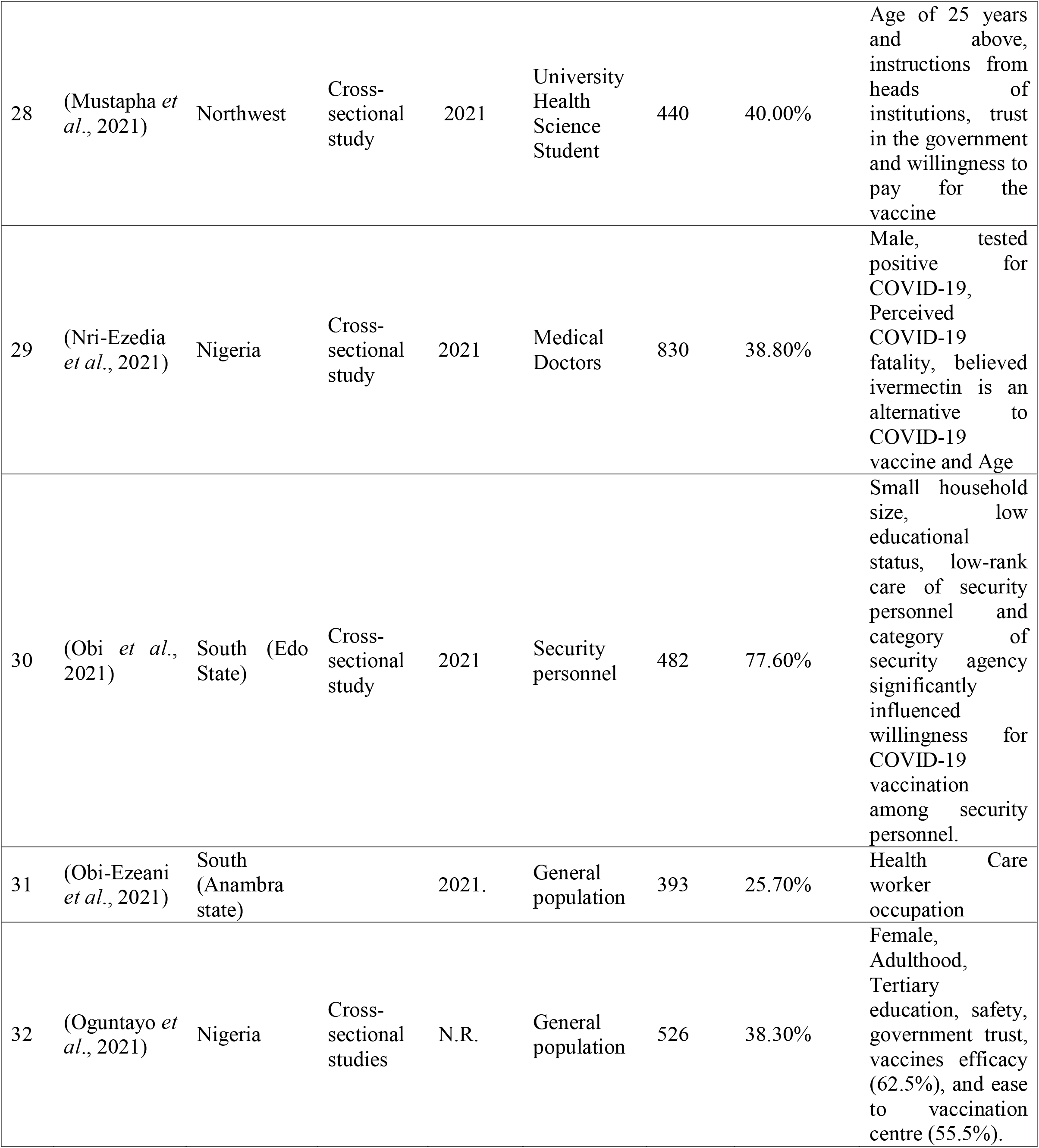
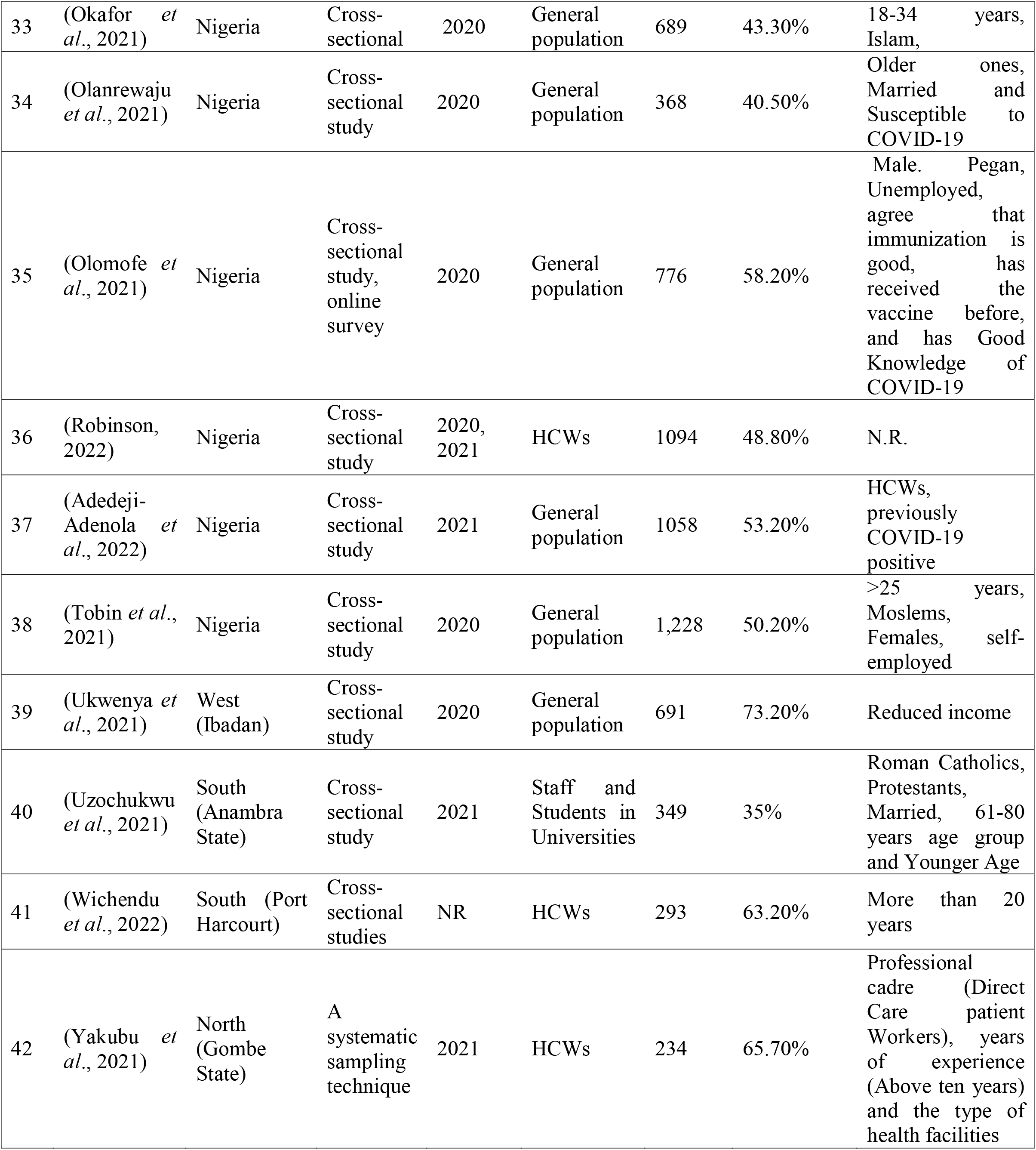
Characteristics of included studies.

### 2.5 Data Analysis

The Meta Prop command was installed in STATA 16 to generate forest plots for vaccine acceptance and hesitancy for the pooled effects. MedCalc was used to create funnel plots for publication bias estimation. Using random-effects models with a 95 % CI, the pooled effects of vaccine acceptance and hesitance were calculated. Begg’s test and Egger weighted regression methods were employed to determine the presence and effect of publication bias.

### 2.6 Assessment of Study Quality

The included papers’ quality was evaluated using the Joanna Briggs Institute (JBI) critical evaluation tool (*Critical Appraisal Tools* | *JBI*, n.d.) Eight questions about the study design and data analysis were included in the checklist (for instance, sample size, sample selection, and reliable and valid measurements). First, all of the scores from each study were added up, and then, based on the results of other studies, they were categorized (Nehal *et al*., 2021; Patwary *et al*., 2022), as shown in *Table 1*.

### 2.7 Search Results

A total of 432 articles were identified in preliminary searches. The Zotero software was used to remove 111 duplications. After eligibility had been assessed based on the title and abstract or the complete text, 71 articles were included in the final selection. Of these, 18 articles were found to lack a precise determination of vaccine acceptance and hesitancy. Ten articles were found inappropriate to include, as 5 were systematic reviews while the other 5 had a qualitative study design. Of the total, 42 studies were included in the meta-analysis *(Figure 1)*.

**Figure 1:**
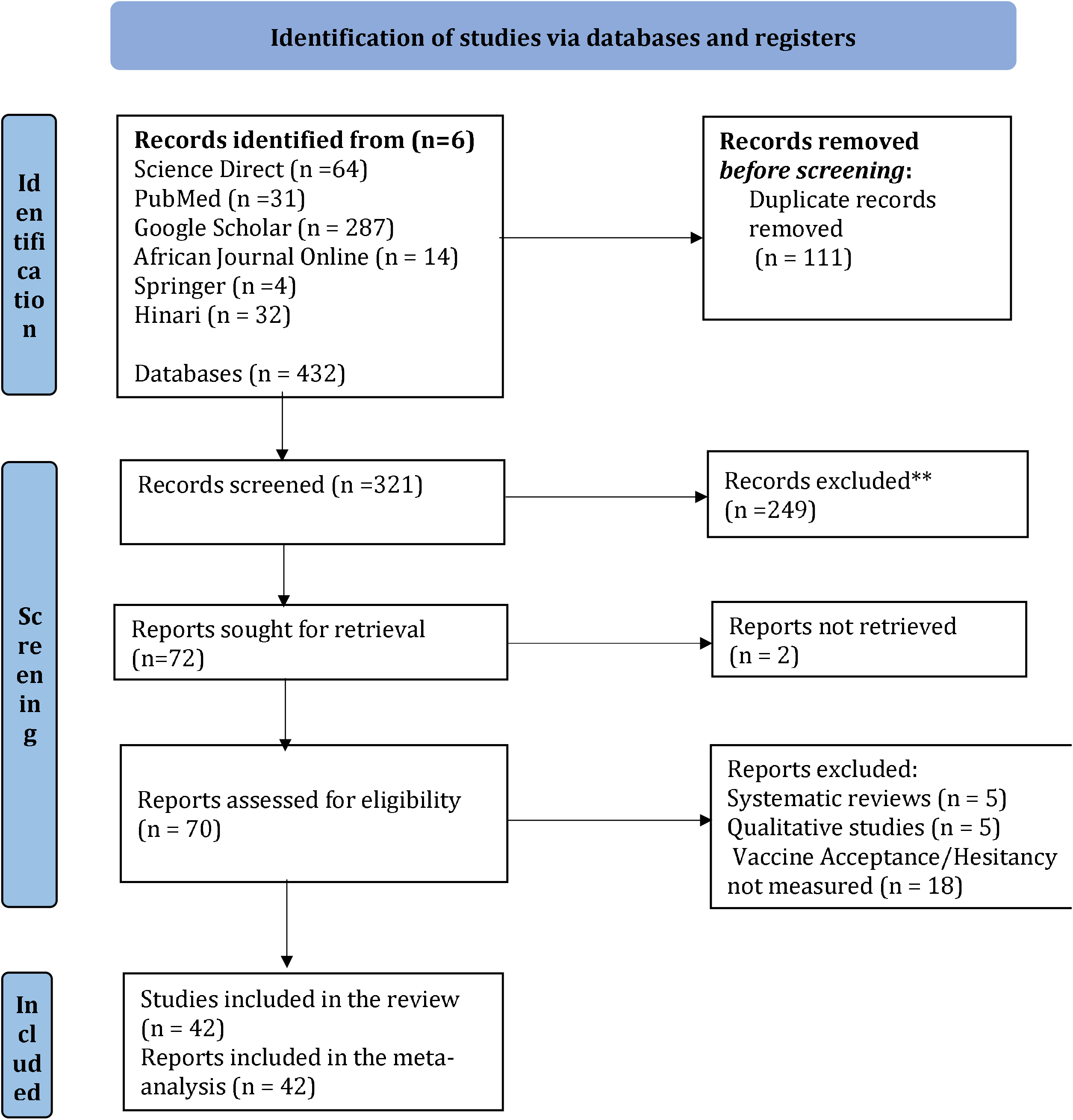
PRISM flow chart

### 2.8 Characteristics of the Included Studies

A brief overview of the included studies and factors affecting vaccine acceptance is shown in Table 1. Most studies used online survey tools and manual questionnaires to collect data for their cross-sectional designs. Few studies used the convenience sampling technique, random sampling, or snowball sampling via email or social media. All surveys were conducted across the board between March 2020 and March 2022. The total sample of included studies was 24,533 ranging from 115 to 1,767 participants in individual studies. The sample size of states in the Northern, Western and Southern parts of Nigeria are 3,206, 4,527 and 5,059, respectively, while 11,741 is the cumulative sample size of all the Nigeria-wide studies.

General populations comprised most of the targeted samples, followed by healthcare professionals and students. The most frequent predictors of vaccine acceptance were older ages, males, marital status, higher education levels, city dwellers, healthcare workers, the presence of chronic diseases, knowledge of COVID-19, perceptions of vaccine risks and benefits, beliefs about the efficacy and safety of vaccines, previous vaccination history, and confidence in healthcare systems.

## 3.0. Prevalence of Vaccine Acceptance and Hesitancy

The total COVID-19 vaccination acceptance rate among all the study groups was 52.4% (95% CI: 46.9-57.9%, *I*^2^ = 100%) (*Figure 2*). In contrast, the total estimated COVID-19 vaccination hesitancy rates were 47.81% (95% CI: 42.2 – 53.4% *I*^*2*^ = 100%) (*Figure 3*). Ilori *et al*., 2021 observed the highest acceptance rate (80.26% CI: 75.46%, 84.3%) in a cross-sectional Nigeria-wide study among Health Care Workers (Ilori *et al*., 2021). Adigwe, 2021 also conducted a similar survey among the general population and reported the lowest vaccination acceptance rate of 22.67% (95%CI 22.80%-24.70%) (Adigwe, 2021).

**Figure 2:**
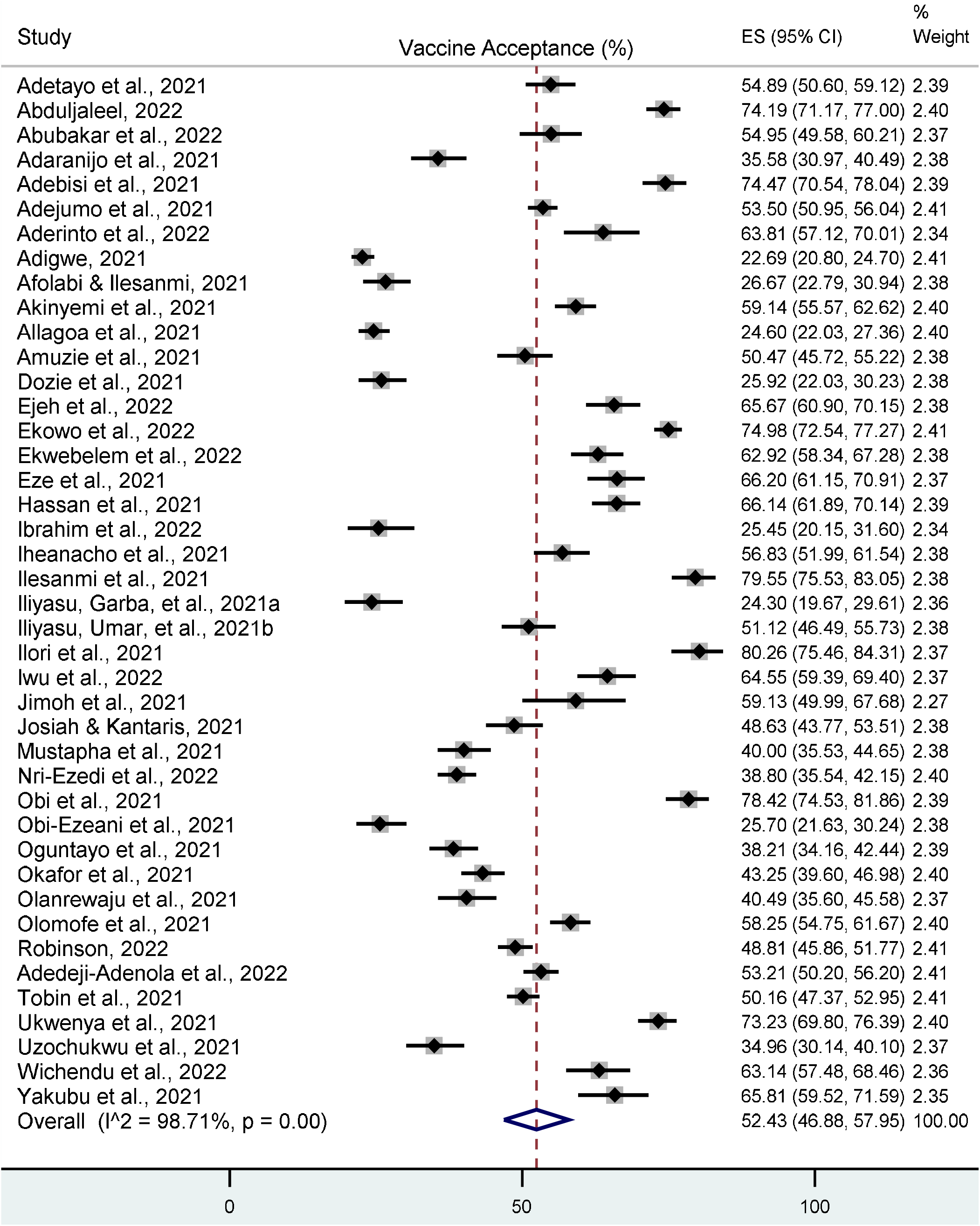
Forest plot of cumulative vaccine acceptance in Nigeria

**Figure 3:**
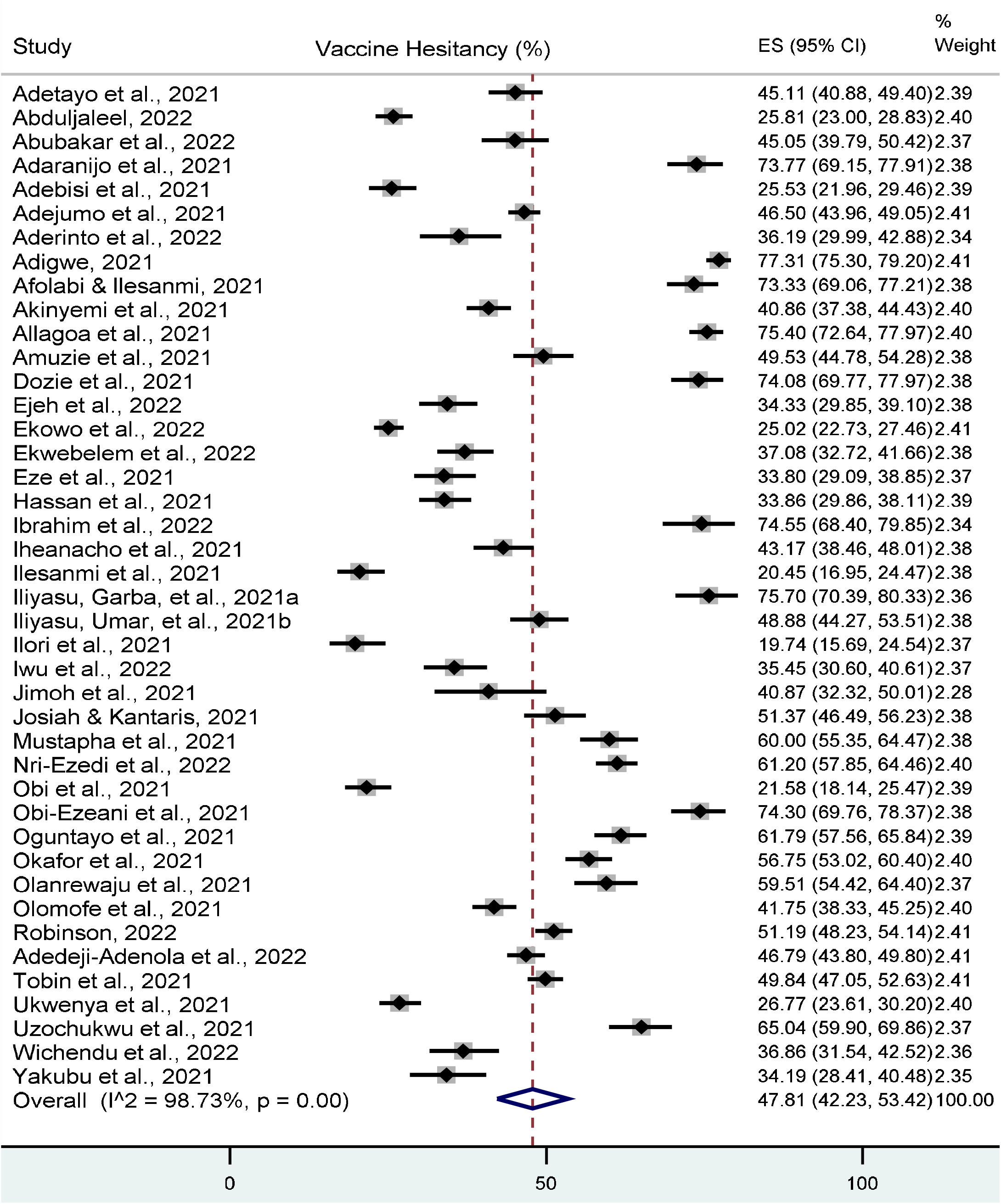
Forest plot of cumulative vaccine hesitancy in Nigeria

### 3.1. Sub-Group Analysis

*Figure 4 and Figure 5* present Western, Northern and Southern regional-specific COVID-19 vaccine acceptance rates and vaccine hesitancy in Nigeria, respectively. The pooled prevalence of the highest acceptance rate was observed in the Western region (58.90%, 95% CI: 47.12– 70.27%), followed by the southern part (47.38%. 95% CI: 31.50–63.55%, *I*^2^ = 99.25%) and Northern region (46.3%, 95% CI: 32.75%–60.18%, *I*^2^ = 98.39%%). Nigeria-wide studies that were randomly sampled, irrespective of regions, were also pooled with a cumulative effect size of vaccine acceptance (55.06%, 95% CI: 47.41%–62.59%).

**Figure 4:**
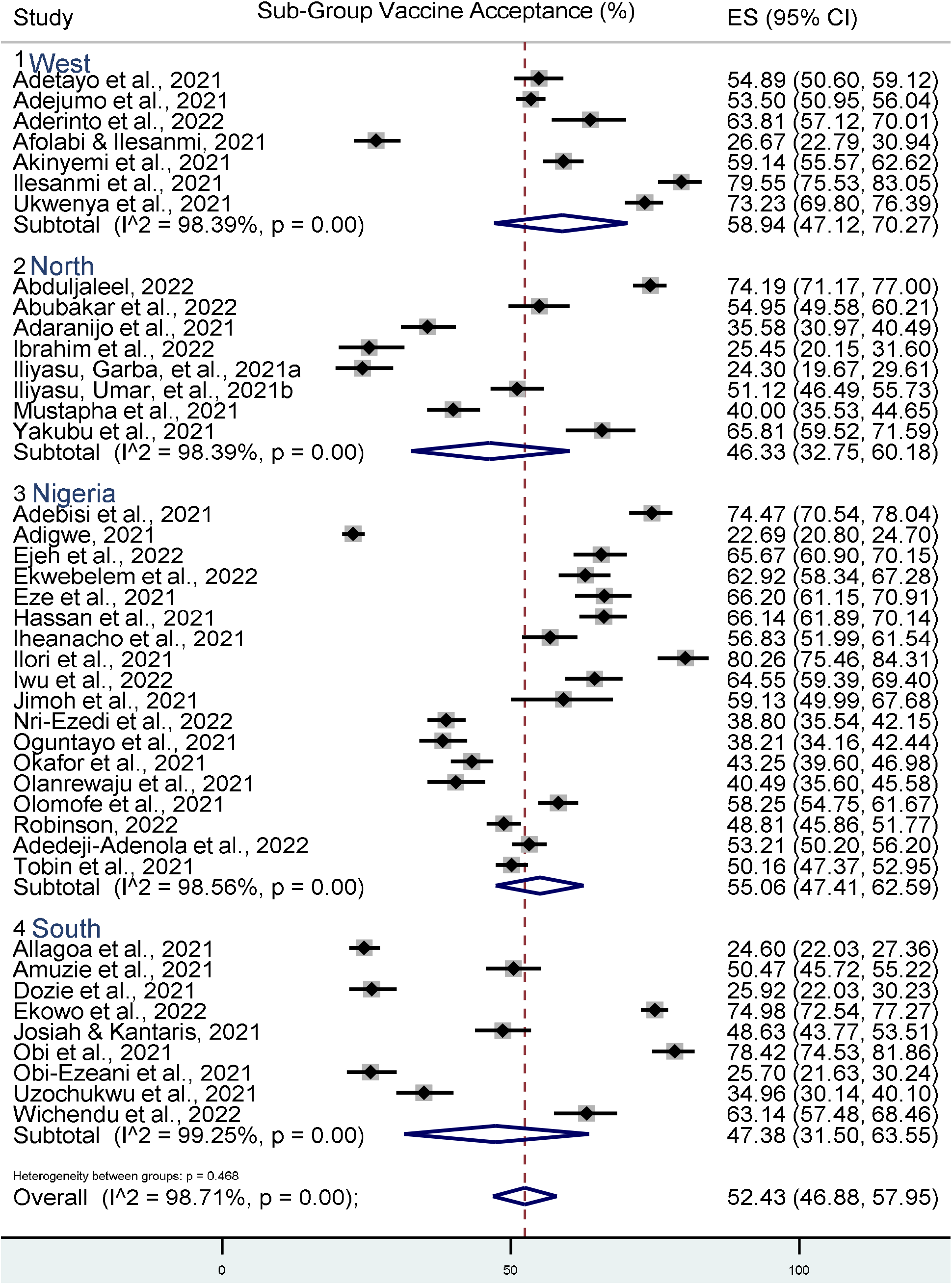
Forest plot of vaccine acceptance in Nigeria across regions

**Figure 5:**
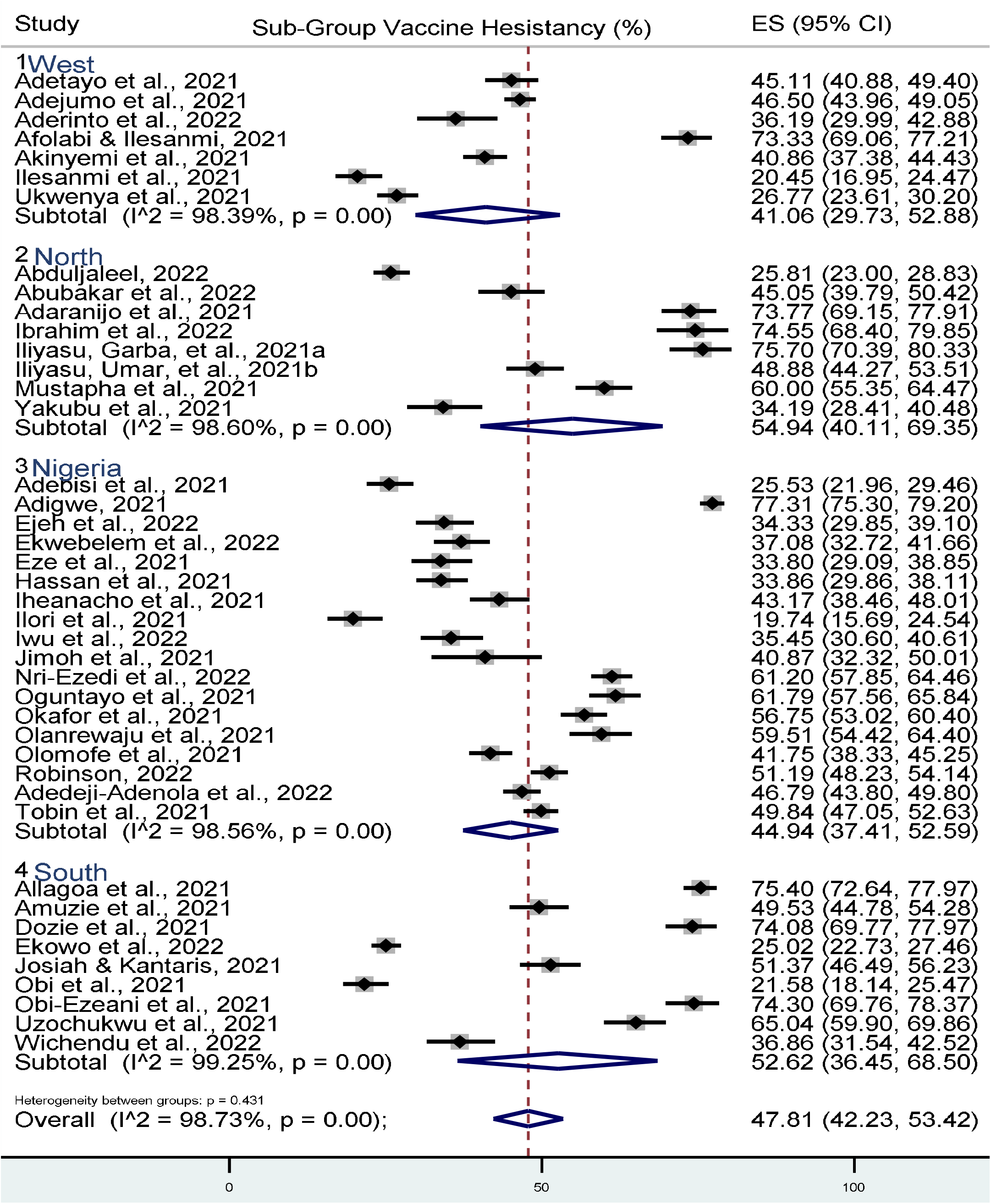
Forest plot of vaccine hesitancy across regions in Nigeria

### 3.2. Time Trends

The COVID-19 vaccine acceptance rate was highest in 2020, with a pooled rate of 59.56% (52.61, 66.32%, *I*^*2*^ = 97.4%). However, the acceptance rate in 2021 was only 48.48 (40.78%, 56.22%), while in 2022, it spiked to 52.04% (95% CI: 35.7%, 68.15 %) (*Figure 6*).

**Figure 6:**
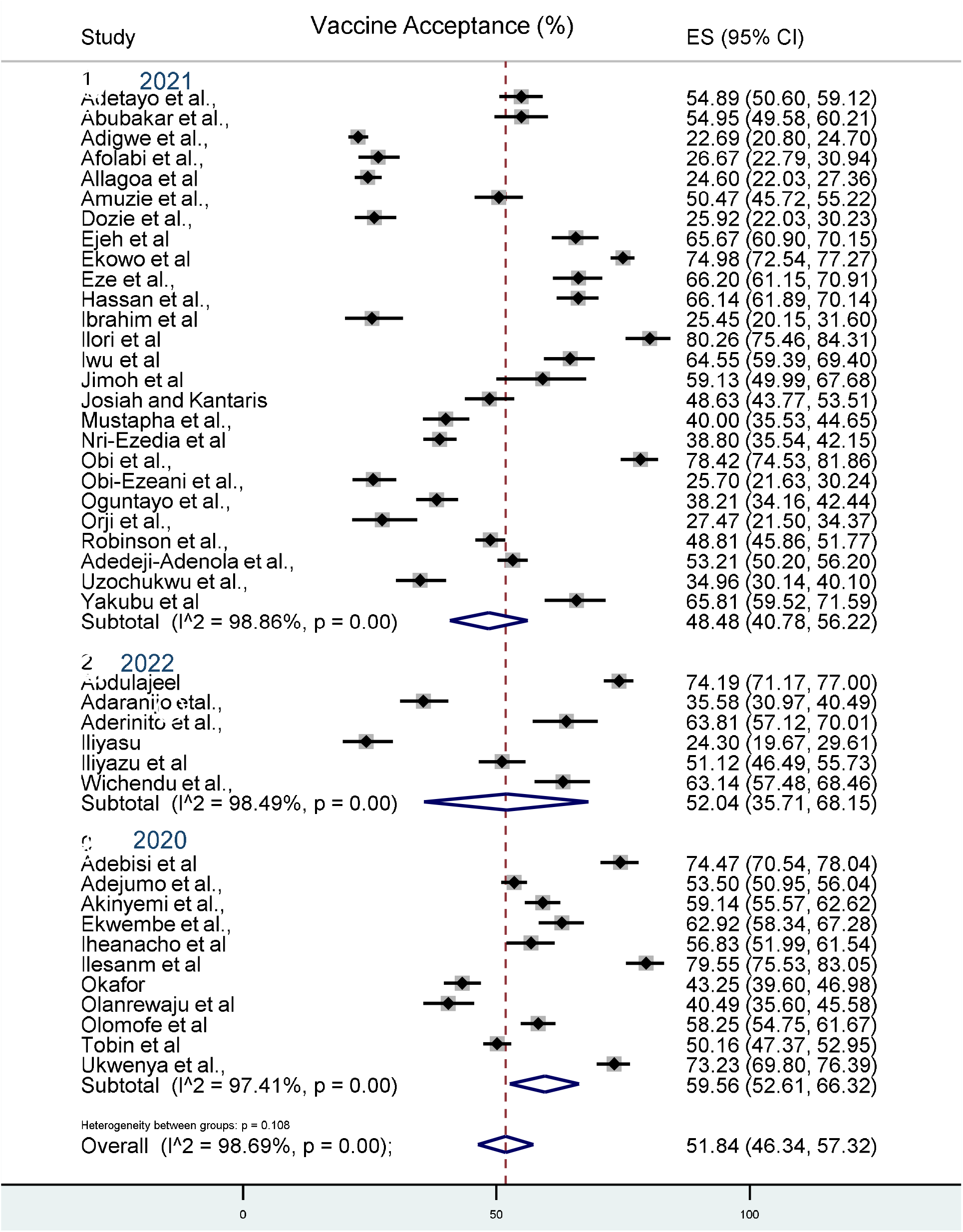
Forest plot of vaccine acceptance across time

### 3.3. Risk of Bias

All 42 studies were assessed to be of the highest possible quality based on the JBI technique (*Table 1*). Studies that used ineffective recruitment methods like convenience and snowball sampling via social media were not removed, but their results may not have been representative of the population. We observed no risk of publication bias. Egger’s tests among studies on vaccine acceptance (*p*-value = 0.4075) and vaccine hesitancy (*p*-value = 0.4437) were not significant (*Tables S1* and *S2*).

## 4. Discussion

### 4.1. The Main Findings in Brief

Several surveys have been conducted since the announcement of efforts to develop a COVID-19 vaccine to measure public perception and acceptance of the vaccine (Pogue *et al*., 2020; Shekhar *et al*., 2021). However, most surveys in Nigeria have concentrated on different states and sub-groups. Therefore, assessing these studies and pooling them together is pertinent to give a representative picture of vaccine acceptance and hesitancy in Nigeria from March 2020 to May 2022. This will assist institutions and policy experts in allocating resources to maximize COVID-19 uptake.

We identified 42 studies of 24,533 participants from various states and geopolitical zones in the country. Pooled estimates showed that more than half (52.4%) of these participants were willing to accept the COVID-19 vaccine. Furthermore, the Southwestern region and Northern Nigeria reported the highest vaccine acceptance and hesitancy rates of 58.9% and 54.9%, respectively, in the sub-group analysis for regions.

The pooled vaccine acceptance in our study is similar to the pooled estimate (58.5%) (Ackah *et al*., 2022; Patwary *et al*., 2022), with 49% on amenability to COVID-19 vaccination in West African countries and low and lower-middle-income countries, respectively. However, higher vaccination acceptance rates in African nations like Nigeria may have been impacted by lower COVID-19 death rates 65. Additionally, there is a history of vaccination scepticism in Africa, which may have affected Nigeria’s poor acceptance rates (Kabamba Nzaji *et al*., 2020).

The relatively higher vaccine acceptance in southwestern Nigeria relative to other parts of Nigeria could be due to the higher risk perception of the pandemic as Lagos state, one of the western states, is the epicenter of the pandemic in Nigeria (Hassan *et al*.,2022). Many studies have demonstrated that increased risk perception of the virus is a decisive predictor variable for COVID-19 Vaccine acceptance (Guidry *et al*., 2021; Lin *et al*., 2021; Shektar *et al*., 2021; Dzieciolowska *et al*., 2021). On the other hand, the high vaccine hesitancy in Northern Nigeria may be related to cultural and religious patterns, which was demonstrated in the past with cases of mass refusal and boycott of the polio vaccination program that led to the resurgence in the diseases and beyond (Ike *et al*., 2018; Nzaji *et al*., 2020; Tobin *et al*., 2021; Adebisi *et al*., 2021). Moreover, the relatively lower figures for the burden of the pandemic in the Northern states of Nigeria could explain the high hesitancy (Hassan *et al*., 2020).

Finally, our study discovered that vaccine acceptability is sinusoidal, declining from 59.56% in 2020 to 48.48 in 2021 before increasing to 52.04% in 2022. The acceptability of vaccines varies throughout time, according to earlier studies (Nehal *et al*., 2021). For instance, a systematic global evaluation of vaccine acceptancy rates found that they had decreased from 79% in March-May to 60% in June–October 2020 (Bono *et al*., 2021). The outcome of our findings may be explained by the fact that Nigerians were more motivated to be vaccinated to prevent infection during the early stages of the epidemic due to their increased level of anxiety. However, according to recent polls (Paul *et al*., n.d.; Roozenbeek *et al*., 2020), the inconsistent trend in vaccination acceptability may be caused by erroneous information and safety worries.

### 4.1. Implications and Future Research

Due to the limited access to healthcare and its dense population, Nigeria is particularly susceptible to COVID-19. Therefore, the Nigerian government should concentrate heavily on achieving high vaccination rates to stop the virus’s spread. In addition, understanding how the general public feels about vaccination is essential for adhering to immunization laws.

This current systematic review and meta-analysis may provide recommendations for subsequent initiatives, given our reported regional and national vaccine uptake and resistance estimations. We advise using state-or region-specific initiatives to increase acceptance rates in Nigeria. The governor of each state should encourage vaccine trust locally in this regard. Governments should be aware of anti-vaccination movements in their states that may be sparked by misunderstanding and disinformation found on social media or from other sources, as these could result in a decline in vaccine acceptance (Danis *et al*., 2010). A better understanding of factors like perceived COVID-19 risk and gender influencing vaccination intentions may also make immunization programs more effective. Given the waves of outbreaks currently occurring in many countries and the fluidity of vaccination acceptance, future research should focus on longitudinal changes in COVID-19 vaccine resistance in Nigeria. In this case, our analysis offers initial insights for interpreting trends in adopting COVID-19 immunization over time. Most of the researched populations in the included studies were drawn from the general community. Future studies should focus on assessing vaccination acceptance rates and determining why some populations, such as healthcare workers, expecting moms, children, and people with chronic conditions, are resistant to immunizations.

### 4.2. Strengths and Limitations

Data collection for our study’s restriction would undoubtedly impact the pooled estimate because COVID-19 vaccination adoption has been observed to fluctuate over time, from March 2020 to May 2022. Additionally, we incorporated a few preprint publications that have not been peer-reviewed. Lastly, we could not assess the predicted drivers for COVID-19 acceptability in Nigeria due to data limitations.

This study has several advantages. It is the first thorough meta-analysis on vaccination acceptability in Nigeria that we are aware of. To provide a typical vaccine acceptability result in Nigeria, we also collected a wide range of research for subgroups and the general population.

## 5.0. Conclusions

In an analysis of 44 papers, we discovered that more than half of Nigerians were receptive to the COVID-19 vaccination. Nigeria’s Western and Northern regions had the highest vaccine acceptance and reluctance levels. Since the pandemic began, 2020 has had the highest level of vaccination acceptance. More studies from the pooled studies revealed that being male and risk perception are frequently linked to vaccine uptake; thus, policymakers at the federal and state levels should consider these factors when developing vaccination policies. The sensitization of local authorities and the dissemination of more detailed information about the COVID-19 vaccine, particularly in the northern, southeastern, and south-central states, could significantly increase the country’s vaccination rate.

## Data Availability

All data produced in the present work are contained in the manuscript

https://drive.google.com/file/d/1ja5dX1wM8lBVTfRMxofwEa5LGdZHZ17q/view?usp=share_link.

## Supplementary Materials

The following supporting information can be downloaded at: https://drive.google.com/file/d/1ja5dX1wM8lBVTfRMxofwEa5LGdZHZ17q/view?usp=share_link.

*Table 2: Characteristics of included studies*,

*Table 2: Joanna Briggs Institute (JBI) critical evaluation table*

*Figure 2: Forest plot of cumulative vaccine acceptance in Nigeria*,

*Figure 3: Forest plot of cumulative vaccine hesitancy in Nigeria*,

*Figure 4: Forest plot of vaccine acceptance in Nigeria across regions*,

*Figure 5: Forest plot of vaccine hesitancy across regions in Nigeria*,

*Figure 6: Forest plot of vaccine acceptance across time*,

*Figure S1: Funnel Plot of Vaccine Acceptance*,

*Figure S2: Funnel Plot of Vaccine Hesitancy*,

*Table S3: Eggers Test on Vaccine Acceptance for publication bias*

*Table S4: Eggers Test on Vaccine Hesitancy for publication bias*

*Table S5: Eggers Test on Vaccine Hesitancy for publication bias*

## REFERENCES

Abduljaleel, A. (2022). Assessment of Knowledge and Acceptance of Covid-19 Vaccinations among Healthcare Workers in Kano State, Nigeria. TEXILA INTERNATIONAL JOURNAL OF ACADEMIC RESEARCH, 9(2), 134–148. https://doi.org/10.21522/TIJAR.2014.09.02.Art011

Abubakar, A. T., Suleiman, K., Idris, A. S., Suleiman, S. Y., Ibrahim, U. B., Abdullahi, S. B., Haladu, A. S., Al-Mustapha, A. I., & Abubakar, M. I. (2022). Acceptance of COVID-19 vaccine among healthcare workers in Katsina state, Northwest Nigeria (p. 2022.03.20.22272677). medRxiv. https://doi.org/10.1101/2022.03.20.22272677

Ackah, M., Ameyaw, L., Salifu, M. G., Asubonteng, D. P. A., Yeboah, C. O., Annor, E. N., Ankapong, E. A. K., & Boakye, H. (2022). COVID-19 vaccine acceptance among health care workers in Africa: A systematic review and meta-analysis. PLOS ONE, 17(5), e0268711. https://doi.org/10.1371/journal.pone.0268711

Adaranijo, E., Badamasi, U., & Haruna, M. (2021). ASSESSMENT OF KNOWLEDGE, PERCEPTION AND PUBLIC ACCEPTANCE OF A COVID-19 VACCINE IN NASARAWA STATE, NIGERIA.

Adebisi, Y. A., Alaran, A. J., Bolarinwa, O. A., Akande-Sholabi, W., & Lucero-Prisno, D. E. (2021). When it is available, will we take it? Social media users’ perception of hypothetical COVID-19 vaccine in Nigeria. The Pan African Medical Journal, 38.

Adedeji-Adenola, H., Olugbake, O. A., & Adeosun, S. A. (2022). Factors influencing COVID-19 vaccine uptake among adults in Nigeria. PloS One, 17(2), e0264371–e0264371. https://doi.org/10.1371/journal.pone.0264371

Adejumo, O. A., Ogundele, O. A., Madubuko, C. R., Oluwafemi, R. O., Okoye, O. C., Okonkwo, K. C., Owolade, S. S., Junaid, O. A., Lawal, O. M., Enikuomehin, A. C., Ntaji, M. I., Sokunbi, A., Timothy, A. O., Abolarin, O. S., Ali, E. O., & Ohaju-Obodo, J. O. (2021). Perceptions of the COVID-19 vaccine and willingness to receive vaccination among health workers in Nigeria. Osong Public Health and Research Perspectives, 12(4), 236–243. https://doi.org/10.24171/j.phrp.2021.0023

Aderinto, N., Afolabi, A., Akande, Y., Ayantoyinbo, O., Samson, A., Ojedokun, S., Bosoro, O., Akatishe, T., & Ilelaboye, A. (2022). Assessment Of Willingness And Acceptability Of Covid-19 Vaccine Among Medical Students In Southwest Nigeria Ayodeji Ilelaboye.

Adetayo, A. J., Sanni, B. A., & Aborisade, M. O. (2021). COVID-19 Vaccine Knowledge, Attitude, and Acceptance among Students in Selected Universities in Nigeria. Dr. Sulaiman Al Habib Medical Journal, 3(4), 162–167. https://doi.org/10.2991/dsahmj.k.211014.001

Adigwe, O. P. (2021). COVID-19 vaccine hesitancy and willingness to pay: Emergent factors from a cross-sectional study in Nigeria. Vaccine: X, 9, 100112. https://doi.org/10.1016/j.jvacx.2021.100112

Afolabi, A. A., & Ilesanmi, O. S. (2021). Addressing COVID-19 vaccine hesitancy: Lessons from the role of community participation in previous vaccination programs. Health Promotion Perspectives, 11(4), 434.

Akinyemi, P., & Akinyemi, P. (2021). Community perception and determinants of willingness to uptake COVID-19 vaccines among residents of Osun State, South-West Nigeria.

Allagoa, D., Oriji, P., Tekenah, E. S., Lukman, O., Chidiebere, N., Sa, A., & Atemie, G. (2021). Predictors of acceptance of COVID-19 vaccine among patients at a tertiary hospital in South-South Nigeria. International Journal of Community Medicine and Public Health, 8, 2165–2172. https://doi.org/10.18203/2394-6040.ijcmph20211733

Amuzie, C. I., Odini, F., Kalu, K. U., Izuka, M., Nwamoh, U., Emma-Ukaegbu, U., & Onyike, G. (2021). COVID-19 vaccine hesitancy among healthcare workers and its socio-demographic determinants in Abia State, Southeastern Nigeria: A cross-sectional study. The Pan African Medical Journal, 40, 10. https://doi.org/10.11604/pamj.2021.40.10.29816

Aw, J., Seng, J. J. B., Seah, S. S. Y., & Low, L. L. (2021). COVID-19 Vaccine Hesitancy-A Scoping Review of Literature in High-Income Countries. Vaccines, 9(8), 900. https://doi.org/10.3390/vaccines9080900

Bono, S. A., Faria de Moura Villela, E., Siau, C. S., Chen, W. S., Pengpid, S., Hasan, M. T., Sessou, P., Ditekemena, J. D., Amodan, B. O., Hosseinipour, M. C., Dolo, H., Siewe Fodjo, J. N., Low, W. Y., & Colebunders, R. (2021). Factors Affecting COVID-19 Vaccine Acceptance: An International Survey among Low- and Middle-Income Countries. Vaccines, 9(5), 515. https://doi.org/10.3390/vaccines9050515

Brown, K., Fraser, G., Ramsay, M., Shanley, R., Cowley, N., van Wijgerden, J., Toff, P., Falconer, M., Hudson, M., Green, J., Kroll, J. S., Vincent, C., & Sevdalis, N. (2011). Attitudinal and demographic predictors of measles-mumps-rubella vaccine (MMR) uptake during the UK catch-up campaign 2008-09: Cross-sectional survey. PloS One, 6(5), e19381. https://doi.org/10.1371/journal.pone.0019381

Critical Appraisal Tools | JBI. (n.d.). Retrieved May 22, 2022, from https://jbi.global/critical-appraisal-tools

Danis, K., Georgakopoulou, T., Stavrou, T., Laggas, D., & Panagiotopoulos, T. (2010). Socioeconomic factors play a more important role in childhood vaccination coverage than parental perceptions: A cross-sectional study in Greece. Vaccine, 28(7), 1861–1869. https://doi.org/10.1016/j.vaccine.2009.11.078

Dempsey, A. F., Schaffer, S., Singer, D., Butchart, A., Davis, M., & Freed, G. L. (2011). Alternative vaccination schedule preferences among parents of young children. Pediatrics, 128(5), 848–856. https://doi.org/10.1542/peds.2011-0400

Ditekemena, J. D., Nkamba, D. M., Mutwadi, A., Mavoko, H. M., Siewe Fodjo, J. N., Luhata, C., Obimpeh, M., Van Hees, S., Nachega, J. B., & Colebunders, R. (2021). COVID-19 Vaccine Acceptance in the Democratic Republic of Congo: A Cross-Sectional Survey. Vaccines, 9(2), 153. https://doi.org/10.3390/vaccines9020153

Dozie, U., Ibe, S., Nwaokoro, C., Chukwuocha, U., Udujih, O., Innocent, C., Dozie, N. S., Nwufo, C., & Onyewu, N. (2021). Compliance with Covid-19 Non-Medicinal Preventive Protocol and Intent to Accept Covid-19 Vaccine Among Adults in South Eastern Nigeria [Preprint]. In Review. https://doi.org/10.21203/rs.3.rs-1083503/v1

Durbach, N. (2000). “They might as well brand us”: Working-class resistance to compulsory vaccination in Victorian England. Social History of Medicine: The Journal of the Society for the Social History of Medicine, 13(1), 45–62. https://doi.org/10.1093/shm/13.1.45

Dzinamarira, T., Nachipo, B., Phiri, B., & Musuka, G. (2021). COVID-19 Vaccine Roll-Out in South Africa and Zimbabwe: Urgent Need to Address Community Preparedness, Fears and Hesitancy. Vaccines, 9(3), 250. https://doi.org/10.3390/vaccines9030250

Ejeh, F. E., Buba, D. M., Reuben, R. C., Jajere, S. M., Denue, B. A., Abunike, S. A., & Malgwi, B. U. (2022). A-One Health Approach on the Evaluation of COVID-19 Risk Perception and Factors Associated with the COVID-19 Vaccine Acceptance in Nigeria, West Africa [Preprint]. In Review. https://doi.org/10.21203/rs.3.rs-1388881/v1

Ekowo, O. E., Manafa, C., Isielu, R. C., Okoli, C. M., Chikodi, I., Onwuasoanya, A. F., Echendu, S. T., Ihedoro, I., Nwabueze, U. D., & Nwoke, O. C. (2022). A cross-sectional regional study looking at the factors responsible for the low COVID-19 vaccination rate in Nigeria. The Pan African Medical Journal, 41, 114. https://doi.org/10.11604/pamj.2022.41.114.30767

Ekwebene, O. C., Obidile, V. C., Azubuike, P. C., Nnamani, C. P., Dankano, N. E., & Egbuniwe, M. C. (2021). COVID-19 Vaccine Knowledge and Acceptability among Healthcare Providers in Nigeria. International Journal of TROPICAL DISEASE & Health, 51–60. https://doi.org/10.9734/ijtdh/2021/v42i530458

Eze, U. A., Ndoh, K. I. N., Ibisola, B. A., Onwuliri, C. D., Osiyemi, A., Ude, N., Chime, A. A., Ogbor, E. O., Alao, A. O., & Abdullahi, A. (2021). Determinants for Acceptance of COVID-19 Vaccine among Nigerians [Preprint]. In Review. https://doi.org/10.21203/rs.3.rs-636090/v1

Gust, D. A., Darling, N., Kennedy, A., & Schwartz, B. (2008). Parents with doubts about vaccines: Which vaccines and reasons why. Pediatrics, 122(4), 718–725. https://doi.org/10.1542/peds.2007-0538

Haby, M. M., Chapman, E., Clark, R., Barreto, J., Reveiz, L., & Lavis, J. N. (2016). What are the best methodologies for rapid reviews of the research evidence for evidence-informed decision making in health policy and practice: A rapid review. Health Research Policy and Systems, 14(1), 83. https://doi.org/10.1186/s12961-016-0155-7

Hassan, Z., Hashim, M. J., & Khan, G. (2020). Population risk factors for COVID-19 deaths in Nigeria at sub-national level. The Pan African Medical Journal, 35(Suppl 2), 131. https://doi.org/10.11604/pamj.supp.2020.35.131.25258

Ibrahim, Z., Ishaq, S. I., & Aliyu, Y. (2022). Acceptance, Knowledge and Attitudes toward COVID-19 Vaccines: A Cross-Sectional Study from Jigawa State, Nigeria. East African Journal of Health and Science, 5(1), 65–72. https://doi.org/10.37284/eajhs.5.1.582

Iheanacho, C., Enechukwu, O. H., & Aguyi-Ikeany, C. N. (2021). Risk Perception of SARS-CoV-2 Infection and Acceptability of a COVID-19 Vaccine in Nigeria [Preprint]. In Review. https://doi.org/10.21203/rs.3.rs-297619/v1

Ike, A., Eleazar, R., Em, M., Nnabuife, O., Nkeonyenasoya, I., Orabueze, A., & Reward, E. (2018). Current Status and Underlying Problems of Eradication of Poliomyelitis in the Remaining Endemic Countries. European Journal of Preventive Medicine, 6. https://doi.org/10.11648/j.ejpm.20180601.15

Ilesanmi, O., Afolabi, A., & Uchendu, O. (2021). The prospective COVID-19 vaccine: Willingness to pay and perception of community members in Ibadan, Nigeria. PeerJ, 9, e11153. https://doi.org/10.7717/peerj.11153

Iliyasu, Z., Garba, M. R., Gajida, A. U., Amole, T. G., Umar, A. A., Abdullahi, H. M., Kwaku, A. A., Salihu, H. M., & Aliyu, M. H. (2022). ‘Why Should I Take the COVID-19 Vaccine after Recovering from the Disease?’ A Mixed-methods Study of Correlates of COVID-19 Vaccine Acceptability among Health Workers in Northern Nigeria. Pathogens and Global Health, 116(4), 254–262. https://doi.org/10.1080/20477724.2021.2011674

Iliyasu, Z., Umar, A. A., Abdullahi, H. M., Kwaku, A. A., Amole, Taiwo. G., Tsiga-Ahmed, F. I., Garba, R. M., Salihu, H. M., & Aliyu, M. H. (2021). “They have produced a vaccine, but we doubt if COVID-19 exists”: Correlates of COVID-19 vaccine acceptability among adults in Kano, Nigeria. Human Vaccines & Immunotherapeutics, 17(11), 4057–4064. https://doi.org/10.1080/21645515.2021.1974796

Ilori, O. R., Ilori, O. S., Awodutire, P. O., Ige, O. R., Idowu, A. B., Balogun, O. S., & Lawal, O. I. (2021). The acceptability and side effects of COVID-19 vaccine among health care workers in Nigeria: A cross-sectional study. F1000Research, 10(873), 873.

Iwu, C. A., Ositadinma, P., Chibiko, V., Madubueze, U., Uwakwe, K., & Oluoha, U. (2022). Prevalence and Predictors of COVID-19 Vaccine Hesitancy among Health Care Workers in Tertiary Health Care Institutions in a Developing Country: A Cross-Sectional Analytical Study. Advances in Public Health, 2022, e7299092. https://doi.org/10.1155/2022/7299092

Jimoh, S. M., Kehinde, O. H., Emmanuel, O. S., & Ahmed, A. (n.d.). Acceptability of Covid-19 Vaccine among Frontline Health Care Workers in North Central and South Western, Nigeria (No. 04). 20(04), 7.

Josiah, B. O., & Kantaris, M. (2021). Perception of Covid-19 and acceptance of vaccination in Delta State Nigeria. The Nigerian Health Journal, 21(2), 60–86.

Kabamba Nzaji, M., Kabamba Ngombe, L., Ngoie Mwamba, G., Banza Ndala, D. B., Mbidi Miema, J., Luhata Lungoyo, C., Lora Mwimba, B., Cikomola Mwana Bene, A., & Mukamba Musenga, E. (2020). Acceptability of Vaccination Against COVID-19 Among Healthcare Workers in the Democratic Republic of the Congo. Pragmatic and Observational Research, 11, 103–109. https://doi.org/10.2147/POR.S271096

Larson, H. J., Cooper, L. Z., Eskola, J., Katz, S. L., & Ratzan, S. (2011). Addressing the vaccine confidence gap. Lancet (London, England), 378(9790), 526–535. https://doi.org/10.1016/S0140-6736(11)60678-8

Larson, H. J., Jarrett, C., Eckersberger, E., Smith, D. M. D., & Paterson, P. (2014). Understanding vaccine hesitancy around vaccines and vaccination from a global perspective: A systematic review of published literature, 2007-2012. Vaccine, 32(19), 2150–2159. https://doi.org/10.1016/j.vaccine.2014.01.081

Luthy, K. E., Beckstrand, R. L., & Peterson, N. E. (2009). Parental hesitation as a factor in delayed childhood immunization. Journal of Pediatric Health Care: Official Publication of National Association of Pediatric Nurse Associates & Practitioners, 23(6), 388–393. https://doi.org/10.1016/j.pedhc.2008.09.006

MacDonald, N. E. & SAGE Working Group on Vaccine Hesitancy. (2015). Vaccine hesitancy: Definition, scope and determinants. Vaccine, 33(34), 4161–4164. https://doi.org/10.1016/j.vaccine.2015.04.036

Mustapha, M., Lawal, B. K., Sha’aban, A., Jatau, A. I., Wada, A. S., Bala, A. A., Mustapha, S., Haruna, A., Musa, A., Ahmad, M. H., Iliyasu, S., Muhammad, S., Mohammed, F. Z., Ahmed, A. D., & Zainal, H. (2021). Factors associated with acceptance of COVID-19 vaccine among University health sciences students in Northwest Nigeria. PLOS ONE, 16(11), e0260672. https://doi.org/10.1371/journal.pone.0260672

Nehal, K. R., Steendam, L. M., Campos Ponce, M., van der Hoeven, M., & Smit, G. S. A. (2021). Worldwide Vaccination Willingness for COVID-19: A Systematic Review and Meta-Analysis. Vaccines, 9(10), 1071. https://doi.org/10.3390/vaccines9101071

Nri-Ezedia, C. A., Okechukwu, C., Ogochukwu, O. C., Nwaneli, E. I., Musa, S., Kida, I. M., Hassan, A. H., & Thomas, U. O. (2021). Predictors of Coronavirus Disease-19 (COVID-19) Vaccine Acceptance Among Nigerian Medical Doctors. Available at SSRN 3820535.

Obi, A. I., Ogbonna, J., Ogaba, M. U., Tobin, E., Okundia, P., Ononigwe, P., Ireye, F., Isara, A. R., & Obi, R. U. (2021). Willingness to accept COVID-19 vaccine and its determinants among selected security personnel in Benin City (No. 1). 2(1), 1–7.

Obi-Ezeani, C., Ilechukwu, O., Onuora, I., Umeaba, G., Onuike, A., Onyegbule, O., Muoneke, N., Nwagbara, I., Nnoruka, O., & Odo, M. (2021). Knowledge, perception and willingness to receive the current COVID-19 vaccine among residents of Awka metropolis, Anambra State, Nigeria. International Journal of Research in Medical Sciences, 9, 3243. https://doi.org/10.18203/2320-6012.ijrms20214405

Oguntayo, R., Olaseni, A. O., & Ogundipe, A. E. (2021). Hesitancy Prevalence and Sociocognitive Barriers to Coronavirus Vaccinations in Nigeria. European Review Of Applied Sociology, 14(23), 24–33. https://doi.org/10.2478/eras-2021-0008

Okafor, U. G., Isah, A., Onuh, J. C., Mgbemena, C. B., & Ubaka, C. M. (2021). Community acceptance and willingness to pay for hypothetical COVID-19 vaccines in a developing country: A web-based nationwide study in Nigeria. The Pan African Medical Journal, 40, 112. https://doi.org/10.11604/pamj.2021.40.112.27780

Olanrewaju, E., Olarinmoye, A., Agofure, O., Akintunde, F., Adebo, O., & Aniyeloye, A. (2021). Willingness to Accept COVID-19 Vaccine and Its Determinants among Nigeria Citizens: A Web-based Cross-sectional Study. Journal of Advances in Medicine and Medical Research, 33, 13–22. https://doi.org/10.9734/JAMMR/2021/v33i830881

Olomofe, C. O., Soyemi, V. K., Udomah, B. F., Owolabi, A. O., Ajumuka, E. E., Igbokwe, C. M., Ashaolu, U. O., Adeyemi, A. O., Aremu-Kasumu, Y. B., Dada, O. F., Ochieze, J. C., Fayemi, O. B., Ologunde, K. W., Popoola, G. O., & Ariyo, O. E. (2021). Predictors of Uptake of a Potential Covid-19 Vaccine Among Nigerian Adults (p. 2020.12.28.20248965). medRxiv. https://doi.org/10.1101/2020.12.28.20248965

Opel, D. J., Robinson, J. D., Heritage, J., Korfiatis, C., Taylor, J. A., & Mangione-Smith, R. (2012). Characterizing providers’ immunization communication practices during health supervision visits with vaccine-hesitant parents: A pilot study. Vaccine, 30(7), 1269–1275. https://doi.org/10.1016/j.vaccine.2011.12.129

Page, M. J., McKenzie, J. E., Bossuyt, P. M., Boutron, I., Hoffmann, T. C., Mulrow, C. D., Shamseer, L., Tetzlaff, J. M., Akl, E. A., Brennan, S. E., Chou, R., Glanville, J., Grimshaw, J. M., Hróbjartsson, A., Lalu, M. M., Li, T., Loder, E. W., Mayo-Wilson, E., McDonald, S., … Moher, D. (2021). The PRISMA 2020 statement: An updated guideline for reporting systematic reviews. BMJ (Clinical Research Ed.), 372, n71. https://doi.org/10.1136/bmj.n71

Patwary, M. M., Alam, M. A., Bardhan, M., Disha, A. S., Haque, M. Z., Billah, S. M., Kabir, M. P., Browning, M. H. E. M., Rahman, M. M., Parsa, A. D., & Kabir, R. (2022). COVID-19 Vaccine Acceptance among Low- and Lower-Middle-Income Countries: A Rapid Systematic Review and Meta-Analysis. Vaccines, 10(3), 427. https://doi.org/10.3390/vaccines10030427

Paudel, S., Palaian, S., Shankar, P. R., & Subedi, N. (2021). Risk Perception and Hesitancy Toward COVID-19 Vaccination Among Healthcare Workers and Staff at a Medical College in Nepal. Risk Management and Healthcare Policy, 14, 2253–2261. https://doi.org/10.2147/RMHP.S310289

Paul, E., Steptoe, A., & Fancourt, D. (n.d.). Anti-vaccine attitudes and risk factors for not agreeing to vaccination against COVID-19 amongst 32,361 UK adults: Implications for public health communications. 21.

Pogue, K., Jensen, J. L., Stancil, C. K., Ferguson, D. G., Hughes, S. J., Mello, E. J., Burgess, R., Berges, B. K., Quaye, A., & Poole, B. D. (2020). Influences on Attitudes Regarding Potential COVID-19 Vaccination in the United States. Vaccines, 8(4), E582. https://doi.org/10.3390/vaccines8040582

Porter, D., & Porter, R. (1988). The politics of prevention: Anti-vaccinationism and public health in nineteenth-century England. Medical History, 32(3), 231–252. https://doi.org/10.1017/s0025727300048225

Robinson. (2022, May 31). Knowledge, acceptance, and hesitancy of COVID-19 vaccine among health care workers in Nigeria. https://www.mgmjms.com/article.asp?issn=2347-7946;year=2021;volume=8;issue=2;spage=102;epage=110;aulast=Robinson

Robison, S. G., Groom, H., & Young, C. (2012). Frequency of alternative immunization schedule use in a metropolitan area. Pediatrics, 130(1), 32–38. https://doi.org/10.1542/peds.2011-3154

Roozenbeek, J., Schneider, C. R., Dryhurst, S., Kerr, J., Freeman, A. L. J., Recchia, G., van der Bles, A. M., & van der Linden, S. (2020). Susceptibility to misinformation about COVID-19 around the world. Royal Society Open Science, 7(10), 201199. https://doi.org/10.1098/rsos.201199

Shekhar, R., Garg, I., Pal, S., Kottewar, S., & Sheikh, A. B. (2021). COVID-19 Vaccine Booster: To Boost or Not to Boost. Infectious Disease Reports, 13(4), 924–929. https://doi.org/10.3390/idr13040084

Tobin, E., Okonofua, M., Adeke, A., & Obi, A. (2021). Willingness to Accept a COVID-19 Vaccine in Nigeria: A Population-based Cross-sectional Study. Central African Journal of Public Health, 7, 53–60. https://doi.org/10.11648/j.cajph.20210702.12

Ukwenya, V., Fuwape, T., Ilesanmi, O., & Afolabi, A. (2021). Willingness to Participate in Testing, Contact Tracing, and Taking the COVID-19 Vaccine among Community Members in a Southwestern State in Nigeria. Global Biosecurity, 3(1), Article 1. https://doi.org/10.31646/gbio.106

Uzochukwu, I. C., Eleje, G. U., Nwankwo, C. H., Chukwuma, G. O., Uzuke, C. A., Uzochukwu, C. E., Mathias, B. A., Okunna, C. S., Asomugha, L. A., & Esimone, C. O. (2021). COVID-19 vaccine hesitancy among staff and students in a Nigerian tertiary educational institution. Therapeutic Advances in Infectious Disease, 8, 20499361211054924.

Wichendu, P., Friday, R., Ijah, R., & Aaron, F. (2022). COVID-19 Vaccine Hesitancy among Health Workers in Surgical Departments in Port Harcourt.

Yakubu, A., Maori, L., G, A., Aliyu, A., Rabiu, I., & A, G. (2021). Study On the Willingness to Accept Covid-19 Vaccine Among Healthcare Workers In Gombe, North-East, Nigeria. European Journal of Pharmaceutical and Medical Research, 8, 775–781.

